# Development of a Conceptual Model of Childhood Asthma to Inform Asthma Prevention Policies

**DOI:** 10.1101/2020.12.15.20248275

**Authors:** Amin Adibi, Stuart E Turvey, Tae Yoon Lee, Malcolm R Sears, Allen B Becker, Piush J Mandhane, Theo J Moraes, Padmaja Subbarao, Mohsen Sadatsafavi

## Abstract

**Background:** There is no definitive cure for asthma; as such, prevention remains a major goal. Decision-analytic models are routinely used to evaluate the value-for-money proposition of interventions. Following best practice standards in decision-analytic modeling, the objective of this study was to solicit expert opinion to develop a concept map for a policy model for primary prevention of asthma.

**Methods:** We reviewed currently available decision-analytic models for asthma prevention. A steering committee of economic modelers, allergists, and respirologists was then convened to draft a conceptual model of pediatric asthma. A modified Delphi method was followed to define the context of the problem at hand (evaluation of asthma prevention strategies) and develop the concept map of the model.

**Results:** Consensus was achieved after three rounds of discussions, followed by concealed voting. In the final conceptual model, asthma diagnosis was based on three domains of lung function, atopy, and their symptoms. The panel recommended several markers for each domain. These domains were in turn affected by several risk factors. The panel clustered all risk factors under three groups of ‘patient characteristic’, ‘family history’, and ‘environmental factors’. To be capable of modeling the interplay among risk factors, the panel recommended the use of microsimulation, with an open-population approach that would enable modeling phased implementation and gradual and incomplete uptake of the intervention.

**Conclusions:** Economic evaluation of childhood interventions for preventing asthma will require modeling of several co-dependent risk factors and multiple domains that affect the diagnosis. The conceptual model can inform the development and validation of a policy model for childhood asthma prevention.

**Funding:** Genome Canada Large-Scale Applied Research Project

## Background

Asthma remains a major cause of morbidity and economic burden across the globe. Asthma is the leading cause of absence from school and the third leading cause of productivity loss.^1–3^ We have estimated the 20-year burden of uncontrolled asthma at $960B in the US, with more than $300B in direct costs alone.^4^ There is no cure for asthma and underdiagnosis in children might lead to permanent lung damage due to tissue remodeling^5^.

The Canadian Healthy Infant Longitudinal Development Study (CHILD)^6^, an ongoing multiethnic birth cohort of 3,455 families launched in 2008, provides an unprecedented opportunity to explore biological and environmental factors that can potentially be modified in early life to prevent asthma. The CHILD team and other investigators have identified antibiotic use, formula feeding, and exposure to phthalates as three modifiable early-life risk factors that can predict asthma diagnosis at the age of five.^7–14^ Emerging evidence suggests a causal relationship between these factors, one that is mediated through either microbiome or immune system, or possibly both.^7–10^

Translation of such knowledge into practice, be it guideline recommendations for risk factor modification or policy-level decisions such as rolling out screening programs, will require careful evaluation of the value of any proposed intervention, which can be quantified in terms of the amount of resources that such interventions consume to produce one unit of health gain. Computer models are an indispensable part of such projections, as long-term outcomes of preventive strategies depend on a tangled network of clinical, economic, and behavioral factors. These decision-analytic models collect evidence from multiple sources and enable projection of intermediate outcomes (e.g., to what extent use of antibiotics during childhood affects the risk of asthma) to policy-relevant metrics (e.g., reduction in costs and gains in Quality-Adjusted Life Years [QALY] with the implementation of a national antibiotic stewardship program).

Conventionally, researchers develop a new decision-analytic model for each specific policy question at hand. However, this fragmented approach is inefficient and leads to unnecessary repetition of model development by many research groups, often with inconsistent assumptions.^15^ This inconsistency makes it hard to compare the results of similar studies. Lack of transparency that comes with insufficient documentation of such *de novo* models makes scientific replication difficult, and ultimately affects the credibility of projections.^16^ Our systematic review of decision-analytic models for asthma interventions concluded that currently-available models are based on inconsistent assumptions and lack the granularity needed to inform ‘Precision Medicine’ policies that take into consideration patient characteristics.^17^ An alternative to such de novo modeling approach is to decouple the model development process from the policy question, thus creating ‘reference models’ that can be used to address multiple policy questions within a unified framework.^15^ This not only increases the consistency across different policy decisions but also enables proper consideration of ‘interactions’ among these decisions. For example, large-scale risk-factor modification attempts for asthma (e.g., reduction in exposure to antibiotics) will change the prevalence of asthma, which in turn affects the yield and cost-effectiveness of pediatric asthma screening programs. Reference models can account for these complex interactions.

As the CHILD study and other global initiatives identify more preventive targets for childhood asthma, we recognize the need for a reference decision-analytic model to evaluate long-term effects of these prevention strategies. An example policy that the model should be able to evaluate would be the use of risk screening tools to identify children at high risk of asthma and modulating gut microbiome to prevent asthma among such high-risk individuals. Another example would be the implementation of a national antibiotic stewardship program towards reducing unnecessary antibiotic exposure in children, thus reducing the risk of asthma.

Here, we review models focused on the primary prevention of asthma and report the formal process through which an expert panel of CHILD study investigators discussed and drafted the concept map for an asthma policy model that reflects the current state of our knowledge about asthma diagnosis.

## Methods

The ISPOR-SMDM Modeling Good Research Practices Task Force has advocated for an explicit process of consulting experts and engaging with stakeholders to convert the problem into the appropriate model structure using influence diagrams and concept maps.^18^ We followed these steps, starting from a scoping review of decision-analytic models of interventions related to primary prevention of asthma to identify any potential models that can be adapted for evaluating asthma prevention strategies in Canada. This was followed by a formal process of consensus building on both the decision problems that the model will have to tackle, and the conceptual structure of such a model in terms of the important domains and associations that need to be considered. The outcome of this exercise was a concept map of an asthma prevention policy model that could guide the subsequent steps of model development.

### Scoping Review

Medline was searched with a combination of MeSH terms *asthma, economic models, cost-benefit analysis, primary prevention*, and *decision support techniques* (details in Appendix I). The search was limited to articles published in English between 2000 to 2020 to reflect recent changes both in our understanding of asthma and the prevalence trends. Abstracts and full texts were reviewed to identify the relevance of the study to the context of asthma prevention and to extract the main features of models used for cost-effectiveness analysis.

### Steering Group and Delphi Panel

We solicited expert opinion on the conceptual relation of early-life factors that lead to asthma through a modified Delphi process. The Delphi process is an established method for achieving consensus among subject experts that encourages equal participation of the panel members through multiple rounds of surveys or interviews.^19,20^ The modified Delphi method that was adopted here involved two rounds of an online survey and a final meeting. Participants had a chance to review and reflect on anonymized survey responses from the first round, before responding to the same questions in the second round. A final meeting was then convened to resolve any remaining disagreements and achieve full consensus, similar to the method previously used for the conceptualization of other health economic models.^21^

A steering group consisting of two economic modelers, an allergist, and two respirologists drafted an initial concept map of childhood asthma, based on the risk factors that the clinical experts deemed relevant in one of the following domains: a risk factor for asthma (e.g., family history), a marker of disease progression (e.g., lung function), or a phenotypic presentation that would affect the likelihood of diagnosis (e.g., symptoms). The first draft of the concept map was based on the findings of the scoping review, but the concept map was expanded to incorporate domains identified by the expert panel. The postulated direction of associations between domains (e.g., lung function affecting symptoms) were identified. We then formed a broader expert panel consisting of six academic clinicians and followed the modified Delphi methodology to refine the concept map and achieve consensus among subject experts.

In the first round of the survey, the participants were asked to 1) evaluate the draft concept map and provide feedback on any missing domain; 2) decide on the direction of associations; 3) rank the strength of each association; and 4) suggest candidate (bio)markers for each domain (details in Appendix II). As recommended, problem and model conceptualization were formulated irrespectively of data availability (that is, factors that need to be modeled should be considered, no matter if there is empirical evidence to populate the model).^18^ In the second round, participants were provided aggregate results of the first round of the survey and asked to answer the same questions after reflecting on their colleagues’ opinions and comments. Results of the second-round survey were shared with panel experts before a final virtual meeting and were discussed to achieve full consensus and finalize the concept map. The surveys were conducted online using Qualtrics and analyzed in Tableau v2020.

### Role of Funding Agencies

The funding agency did not have a role in the design and analysis of this study.

## RESULTS

### Scoping Review

Our search strategy resulted in 11 indexed abstracts published between 2000 to 2020. We reviewed the abstracts and excluded 5 citations for not including a decision mode (n=4), and a language other than English (n=1). The remaining citations (n=6) were fully reviewed. Of these, two were excluded for not targeting asthma prevention as the primary outcome. The remaining 4 studies were included in the analysis.^22–25^ ***Figure 1*** shows the flow of the studies in the scoping review. ***Table 1*** summarizes the included studies. All identified models were simple decision-trees or Markov models designed to address the cost-effectiveness of a single intervention.^22–25^

**Table 1.** Summary of scoping review of primary prevention modeling studies for asthma.

| Study | Model | Time-horizon | Perspective | Population | Intervention | Comparator |
| --- | --- | --- | --- | --- | --- | --- |
| Yieh (2019)<sup>22</sup> | Decision-tree | Birth to age 18 | Healthcare and societal | Offspring of pregnant smokers | Vitamin C supplement | Standard prenatal vitamin |
| Ditkowsky (2017)<sup>23</sup> | Markov | Birth to age 20 | Societal | US population | Varicella Vaccination | No universal varicella vaccination |
| Ramos (2014)<sup>24</sup> | Decision-tree | Birth to age 6 | Healthcare system | Unborn children | Uni and multi-faceted primary prevention | Usual care |
| Ramos (2011)<sup>25</sup> | Decision-tree | Birth to age 6 | Healthcare system | Unborn children | Uni and multi-faceted primary prevention | Usual care |

**Figure 1.**
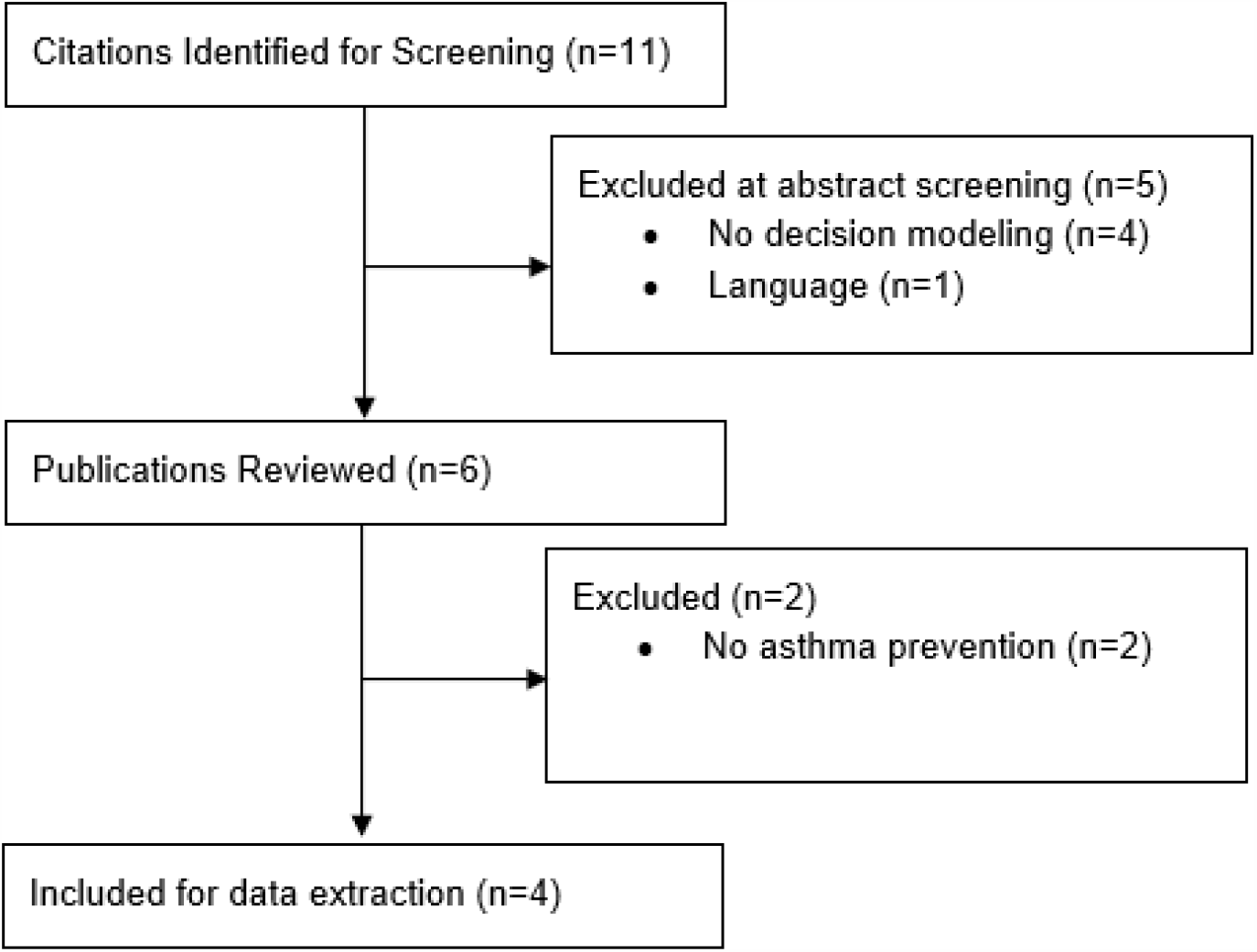
Flow of analysis for the systematic review.

We identified only one publication focused on the conceptualization of an asthma prevention model. Ramos and colleagues reported structuring and validating of a cost-effectiveness model of primary asthma prevention through allergen avoidance.^25^ The authors developed the structure of the model through round-tables and validated it via further discussions with experts and comparison to other asthma models.

We did not identify any reference models for the primary prevention of pediatric asthma. Our conclusion from the scoping review was that none of the previous models can be adapted to act as a reference policy model for childhood asthma prevention.

### Conceptualization of the Decision Problem

Discussions among experts led to the decision problem framework shown in ***Table 2***. The panel decided that the objective of the policy model should be to evaluate the cost-effectiveness of asthma prevention strategies in the pediatric population living in urban and rural settings in Canada. Outcomes of interest were the incidence and prevalence of asthma, asthma-related hospital admissions, direct and indirect costs, and QALYs for patients and their caregivers from a societal perspective. The group decided not to model constrained resources such as access to specialist physicians for the first version of the model.

**Table 2.** Objectives, scope, and policy context of primary prevention model of asthma in children.

| Framework Aspect | Details |
| --- | --- |
| Policy context | To evaluate cost-effectiveness of asthma prevention strategies |
| Funding source | Genome Canada |
| Disease | Childhood asthma |
| Perspective | Societal |
| Target population | Pediatric population in urban and rural settings of Canada |
| Health outcomes | Incidence and prevalence of asthma, asthma-related admissions, and QALY effects on caregivers |
| Strategies/comparators | Current standard of care vs asthma risk screening tool (early diagnosis) with any combination of preventative strategies (e.g. modulating gut microbiome, reducing unnecessary antibiotics/phthalate exposure) |
| Cost(s) | Costs from a societal perspective |
| Time-Horizon | 2025-2045 |

The target of this consensus making exercise was to conceptualize the natural history of asthma up to diagnosis, as the natural history after diagnosis has been extensively modeled in the literature, as shown in our previous systematic review.^17^

### Conceptualization of the Model

The initial draft of the concept map which was developed by the steering group related asthma diagnosis to biological sex, ethnicity, environment, genetics, birth mode, breastfeeding, and infections. The effects were considered to be modulated through lung function, atopy, and possibly microbiome, as shown in ***Figure 2A***.

**Figure 2.**
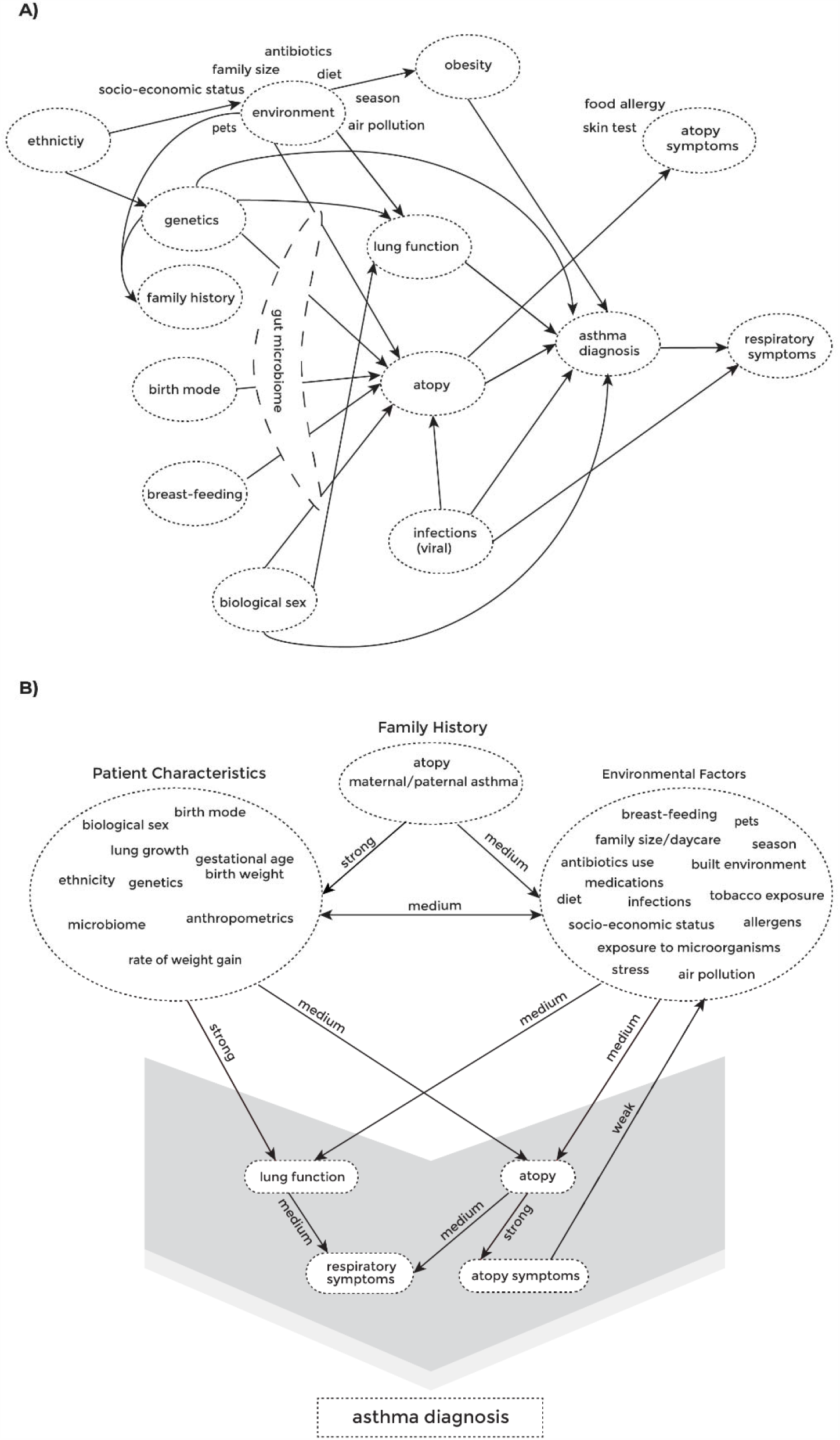
Initial (A) and final (B) concept maps for asthma diagnosis.

The Delphi panel refined the draft concept map through two rounds of surveys and a final consensus meeting. Results of the survey are summarized in Appendix III. The panel added additional factors, renamed some factors for clarity, and clustered all factors under three domains of ‘patient characteristic’, ‘family history’, and ‘environmental factors’. The panel reviewed the direction of causality and assigned strength of relationship to each line in the concept map, as shown in ***Figure 2B***. The final concept map indicated that the likelihood of a diagnosis of asthma in a child is influenced by multiple factors, namely respiratory symptoms, lung function, and atopy. The panel proposed a list of potential measures for these factors, as shown in ***Table 3***.

**Table 3.** Proposed markers of the disease.

| Concept | Measures |
| --- | --- |
| Lung Function | Normalized spirometry measure, bronchoprovocation measures, Reversibility |
| Atopy | Immune trajectory, aeroallergen/food through in-vivo or in-vitro testing |
| Atopy Symptoms | Allergic rhinoconjunctivitis, atopic dermatitis, allergic rhinitis, food allergy |
| Respiratory Symptoms | Wheeze, cough, episodic shortness of breath |

Given the multidimensionality of risk factors, it was decided that the unit of representation in the model will be individual subjects, followed from their date of birth (i.e., microsimulation). The team recommended a discrete event simulation approach, but also agreed that a discrete time approach with weekly or monthly cycles would also be an acceptable alternative. Microsimulation was deemed particularly advantageous for building a reference model, given its ability to capture complex interactions among decisions. Further, the team recommended that the model should follow an open population, due to the importance of modeling realistic aspects of rolling out an intervention or policy, such as the phased implementation of a national screening program. Given the open population nature of the model, the time horizon would be on the calendar year, instead of up to a certain patient age commonly used in previous studies. The team also decided that the model should be open-source and easily accessible to the wider research community.

## DISCUSSION

Economic evaluation of asthma prevention interventions is timely, particularly because of the recent identification of early life exposures that might be causally related to asthma diagnosis later in life.^7–14^ Any asthma prevention strategy would require either population-level or individual-level modification of early-life risk factors that impact the disease.^26^ Recognized risk factors range from those that might be modifiable at the patient-level (e.g. birth mode, breast-feeding, diet, pets, antibiotics, tobacco), those that might be modifiable but require change at the societal level (e.g., air pollution, socio-economic status), and those that cannot be modified (e.g. biological sex, family history, genetics). Some strategies, such as reducing unnecessary antibiotics exposure, can – and perhaps should – be adopted across the board. Other potential preventative interventions (such as modification of the microbiome) may be more suitable for high risk individuals given their cost and possible side-effects.

For clinicians, the decision problem is two-fold: who is at high risk for developing asthma and what intervention has the highest preventive potential for the individual? Both at the societal level and from the perspective of the health-care system, the question becomes what combination of screening and risk-reduction strategies is cost-effective for asthma prevention. Our ultimate goal is to develop a modeling platform for the evaluation of the value-for-money potential of such interventions and policies in a unified framework. Here, we solicited expert opinion and developed a concept map for a reference model that can assess the cost-effectiveness of any combination of risk prediction and prevention strategies.

The ISPOR-SMDM task force highlights the risk of payer’s influence on the analysis.^18^ An independently funded reference model designed to address a variety of policy questions will mitigate such risk. By decoupling model development from any particular policy question, we have eliminated the risk of adopting assumptions that may favor the sponsor’s outcome of interest.^27^ Further, the open-source and accessible nature of the model will facilitate independent evaluation and validation of the model.

Our systematic review of primary prevention models for asthma identified one model conceptualization^25^ and 3 modeling publications;^22–24^ all four were Markov or decision-tree models that modeled a single preventative intervention as a lump-sum reduction in the probability of asthma diagnosis. None modeled the underlying disease processes. In contrast, our conceptual model is based on the progression of personal and environmental risk factors, as well as atopy, lung function, and symptoms over time. Our model structure also accounts for the interplay between these factors such as modification of lifestyle and environmental risk factors in response to worsening symptoms.

In summary, we have followed a formal procedure to solicit expert opinion and draft a concept map for an asthma prevention model. The conceptual model will form the basis for the development and validation of a microsimulation policy model for childhood asthma prevention. This level of granularity allows for evaluating cost-effectiveness analysis of a wide range of screening and intervention methods. For example, the resulting model can be used to inform a precision-medicine approach towards asthma prevention based on considering multiple risk factors (e.g., using a validated risk prediction model). The model will be capable of identifying the optimal risk threshold that would justify preventive interventions, and the optimal preventive intervention for individuals with a given risk profile. With the identification of a growing number of potentially modifiable risk factors, and the advent of novel preventive strategies, the need for exploring such a complex decision space is becoming more pressing.

## Data Availability

All survey data is available upon request.

## ACKNOWLEDGMENTS

We would like to thank Respiratory Evaluation Sciences Program team at the University of British Columbia for their input and feedback and Zahra Jalali for helping with the graphics.

