## Supplementary Material for "Development of a Conceptual Model of Childhood Asthma to Inform Asthma Prevention Policies"

### Appendix I: Search Strategy for the Systematic Review

Medline was searched with the MeSH term below.

"((Asthma/economics[Mesh]) AND Primary Prevention[Mesh]) OR (Primary Prevention[Mesh] AND Asthma[Mesh] AND Models, Economic[Mesh]) OR (Asthma[Mesh] AND Cost-Benefit Analysis[Mesh] AND Primary Prevention[Mesh]) OR (Primary Prevention[Mesh] AND Asthma[Mesh] AND Decision Support Techniques[Mesh])" .

Results were up to date as of December 2020.

Table S1 – Citations found during the Literature Search

| Citation | Journal | Screening Review | Full Paper Review |
| --- | --- | --- | --- |
| Yieh (2019) | J Perinatol | Included | Included |
| Ditkowsky (2017) | Pediatr. Infect. Dis. J. | Included | Included |
| Comert (2014) | J Asthma | Excluded; No modelling. | --- |
| Ramos (2015) | Eur J Health Econ | Included | Included |
| Ramos (2012) | BMC Med Res Methodol | Included | Included |
| Lenoir-Wijnkoop (2012) | Eur J Health Econ | Included | Excluded. Asthma not the primary outcome |
| Bueving (2007) | Curr Allergy Asthma Rep | Included | Excluded. No asthma prevention. No modeling. |
| Pandalis (2006) | MMW Fortschr Med | Excluded; Language. | |
| Kozma (2004) | Int. Immunol. | Excluded; No modelling. | |
| Custovic (2001) | Clin Rev Allergy Immunol | Excluded; No modelling. | |
| van den Boom (2000) | Prev Med | Excluded; No modelling |  |
| Heinen (1994) | Manag Care Q | Excluded; too old |  |

### Appendix II: Survey Questions

1. Arrows shown in the concept map (above) represent causal relationships.  Are there any concepts (nodes) missing in the map that should be added?
   a. Recommend the most appropriate measure for each of the attributes below:
      Asthma, Atopy Symptoms, Atopy, Lung Function, Respiratory Symptoms
   b. Are there any relationships (lines) or directions that are missing in the concept map shown?
2. To make the plot simpler and more understandable, we have clustered patient characteristics and environmental factors together, as shown above.
   a. Do you agree with the way items are clustered? If not, use the form below to  drag and drop items between the clusters.
   b. Are there any important variables missing within the patient characteristics cluster? Please name them and specify which cluster they belong to.
   c. Are there any important variables missing within the environmental variables cluster? Please name them and specify which cluster they belong to.
3. In this part of the survey, we ask for your expert opinion about relationships between concepts (lines that connect the nodes). For each question, we highlight a line, and ask you to comment on whether the direction of the relationship is correct. We also ask you to qualitatively rank the strength of the relationship.

Is the direction of the relationship between patient characteristics and family history (the red arrow) correct?

a. How would you rank the strength of the relationship between patient characteristics and family history (the red arrow)?
b. Q133 please elaborate on your answer if needed.
4. Is the direction of the relationship between patient characteristics and atopy (the red arrow) correct?
   a. How would you rank the strength of the relationship between patient characteristics and atopy (the red arrow)?
   b. please elaborate on your answer if needed.
5. Is the direction of the relationship between patient characteristics and lung function (the red arrow) correct?
   a. How would you rank the strength of the relationship between patient characteristics and lung function (the red arrow)?
   b. please elaborate on your answer if needed.
6. Is the direction of the relationship between environmental factors and family history (the red arrow) correct?
   a. How would you rank the strength of the relationship between environmental factors and family history (the red arrow)?
   b. please elaborate on your answer if needed.
7. Is the direction of the relationship between environmental factors and atopy (the red arrow) correct?
   a. How would you rank the strength of the relationship between environmental factors and atopy (the red arrow)?
   b. please elaborate on your answer, if needed.

8. Is the direction of the relationship between environmental factors and lung function (the red arrow) correct?
   a. How would you rank the strength of the relationship between environmental factors and lung function (the red arrow)?
   b. please elaborate on your answer, if needed.
9. Is the direction of the relationship between lung function and respiratory symptoms (the red arrow) correct?
   a. How would you rank the strength of the relationship between lung function and respiratory symptoms (the red arrow)?
   b. please elaborate on your answer, if needed.
10. Looking back at the concept map, are there any concepts (nodes) missing in the map that should be added? Are there any relationships (lines) or directions that are missing in the concept map shown?

### Appendix III: Summary of Survey Results

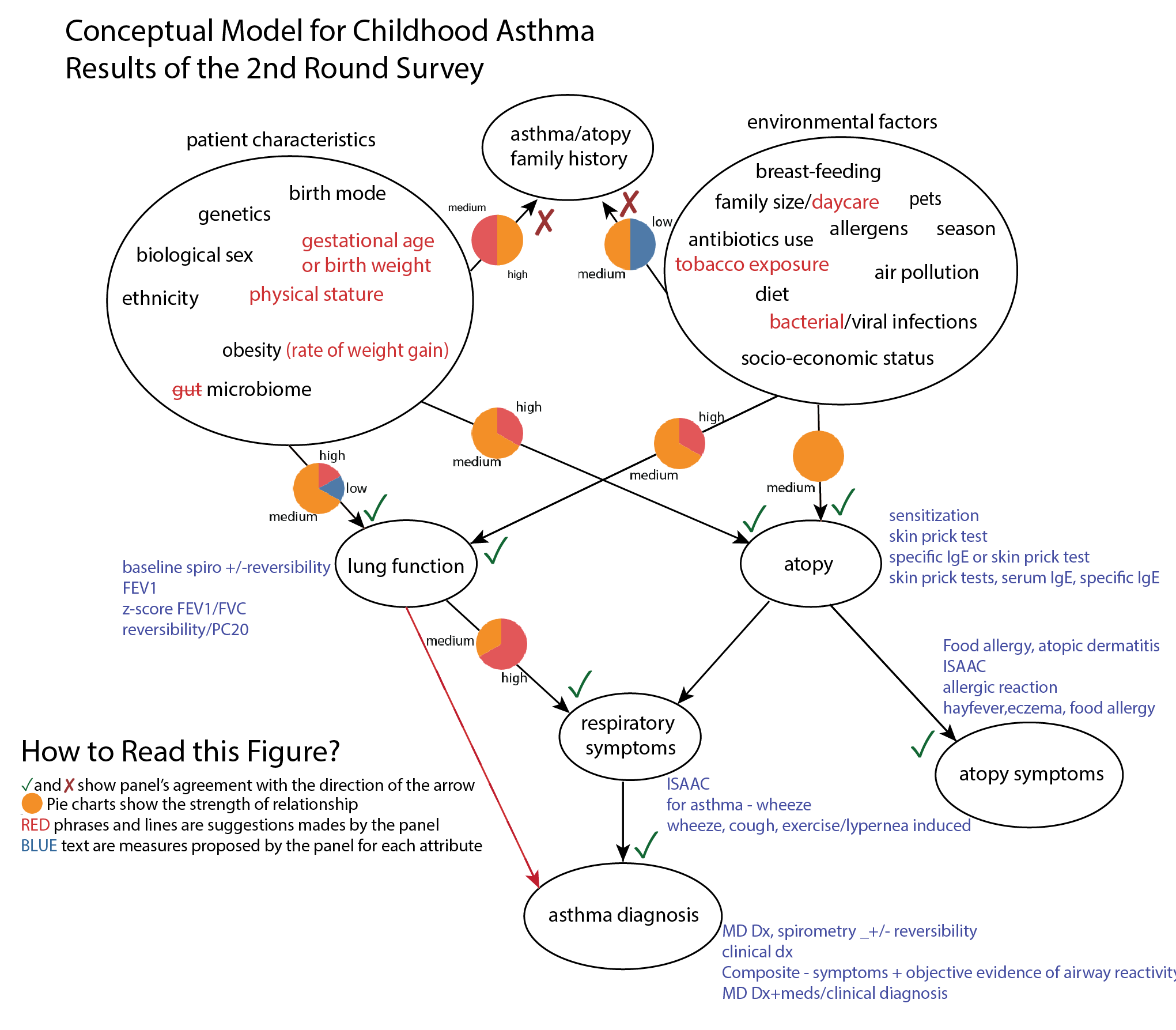
